# The assessment of family experience and expected outcomes from early intervention in preschool children with Autism Spectrum Disorder: translation and cross-cultural adaptation of the Autism Family Experience Questionnaire (AFEQ) to the Italian context

**DOI:** 10.1101/2024.08.16.24311812

**Authors:** Rocchi Claudia, Bonetti Francesca

## Abstract

**Background:** Autism spectrum disorder (ASD) is a neurodevelopmental disorder characterized by deficits in social communication and the presence of restricted interests and repetitive behaviours (Hodges et al., 2020). According to the American guidelines, the impact of disability can affect the family’s well-being and have consequences for the siblings and the organisation of family life, both economically and psychologically (ASHA, 2006). Given the lack of parent-focused measures to assess these aspects, Leadbitter et al. (2018) developed the Autism Family Experience Questionnaire (AFEQ), aimed at assessing priority outcomes for parents of preschool-aged children with ASD who were previously enrolled in the Pre-school Autism Communication Trial, a randomized controlled trial study on parent-mediated intervention (Leadbitter et al., 2018; Green et al., 2010). The aim of the present study was to translate and culturally adapt into Italian the AFEQ (Leadbitter et al., 2018) and to verify the main psychometric performances of the adapted version.

**Methods:** Linguistic validation and cross-cultural adaptation to the Italian context were based on the guidelines of Beaton et al. (2000). Once the direct and inverse translation phases were completed with the favourable opinion of the panel of experts recruited for the study, further tests on the psychometric properties of the adapted questionnaire were required. Following the consensus obtained from the authors of the original questionnaire and according to the recommendations of Polit et al. (2007) and Polit and Beck (2006), a second panel of selected experts determined the content validity of the adapted instrument through the calculation of the Content Validity Index (CVI), according to the indications for each item (I-CVI) and for the scale (S-CVI).

**Results:** A medical doctor and a non-medical expert, both native Italian speakers with an adequate knowledge of English, independently produced a translation. The latter were then compared and discrepancies in the translation process were resolved in a discussion. Two independent, native English-speaking translators produced reverse translations of the resulting version; neither of them had experience or knowledge of the objectives of the study nor of the healthcare sector. The two translations were similar to each other and one of them was almost identical to the original. The original questionnaire and the material obtained in the previous stages were made available to an interdisciplinary panel of experts recruited for the study. All members agreed on that T1-2 was the only accepted translated version. Analysing the I-CVI, all items exceeded the expected cut-off of ≥ 0.78 except for one (I-CVI value 0.6). The S-CVI reached and exceeded the cut-off when obtained with the S-CVI/Ave calculation mode, while with the S-CVI/UA the value was 0.73 and was slightly below the cut-off.

**Discussion and Conclusion:** To the authors’ knowledge, this study is the first to develop an Italian version of the AFEQ questionnaire. The involvement of two translators in the forward and reverse translation phases avoided, in the former, the presence of bias and the enrichment of the synthesis process; in the latter, it facilitated the verification of the semantic equivalence of the translation. Evaluation of the final version by a panel of experts ensured its linguistic validity. The Italian version of the AFEQ obtains very positive I-CVI, S-CVI/UA-Ave values, meaning a very positive assessment of the scale items regarding their relevance for the investigated constructs. As the study is still in progress, it has not yet been possible to carry out a test of the adapted version that coincides with the final phase of the adaptation process described by Beaton et al. (2000). The work represents a first step in the Italian validation process of the questionnaire, which is still ongoing.

## Introduction

In the past few years, among the priorities of the national health services is to be able to measure and monitor the outcomes of the rehabilitation interventions provided, especially those outcomes that are perceived as a priority by patients (Morris et al., 2015). To be meaningful, the outcomes measured should not only meet patients’ and caregivers’ expectations, but also be consistent with the goals that healthcare professionals hope to achieve for the patient (Morris et al., 2015). However, this vision is not always shared by the three parties involved (Kennedy, 2010), which makes it necessary to agree on goals in advance and to establish tools and measures to assess their achievement (Morris et al., 2015). In the research conducted by Morris and colleagues (2015) it emerges that for families and caregivers the appropriate measures for assessing the outcome of rehabilitation intervention in children and young people with Neurodevelopmental Disorders include: communication, emotional well-being and mental health, pain, sleep, mobility and independence, self-care, social life, behaviour and safety.

When evaluating the outcomes of interventions targeting young children with ASD, in addition to the child’s quality of life, it is recommended to follow the broadest possible approach that also includes the well-being of the parents and the entire family system as an important outcome within autism interventions (Leadbitter et al, 2018); furthermore, despite the fact that parents of children with ASD are often the most likely to observe and report on their children’s progress, especially when their children are young or lack the capacity to do so (e.g. children with intellectual or communication disabilities), there are few tools in the literature that are aimed at them and that set out to assess the achievement of intervention outcomes for children with Autism Spectrum Disorder from the perspective of their families (Leadbitter et al., 2018). As pointed out by Dr Leadbitter and her research team, aspects of mental health (e.g. parental stress) or quality-of-life indicators specific to parents of children with ASD are commonly used to survey the well-being of the parents of these children, whereas there was no measure of family experience in the literature that could be used to assess the impact of an intervention in children with ASD.

The Autism Family Experience Questionnaire - AFEQ was therefore created with the intention of formulating a measure aimed at parents of children with ASD that assesses family experience, quality of life and outcomes of early rehabilitation intervention deemed important to the family (Leadbitter et al., 2018). The Questionnaire was developed and tested as part of a large, randomised trial of a parent-mediated intervention targeting pre-school children (Leadbitter et al., 2018), Pre-school Autism Communication Therapy - PACT Therapy (Green et al., 2010). The AFEQ, the characteristics of which are described in Table 1, has been shown to be an ecologically valid instrument with good internal consistency, easy to fill out by parents of children with ASD, sensitive to intervention outcomes and change; for these reasons it can be considered a valid and useful tool, both in research and clinical practice (Leadbitter et al., 2018). The aim of the present work is to conduct the linguistic validation and cross-cultural adaptation of the Autism Family Experience Questionnaire (AFEQ) to the Italian context as a first step in the validation process of the instrument.

**Table 1.**
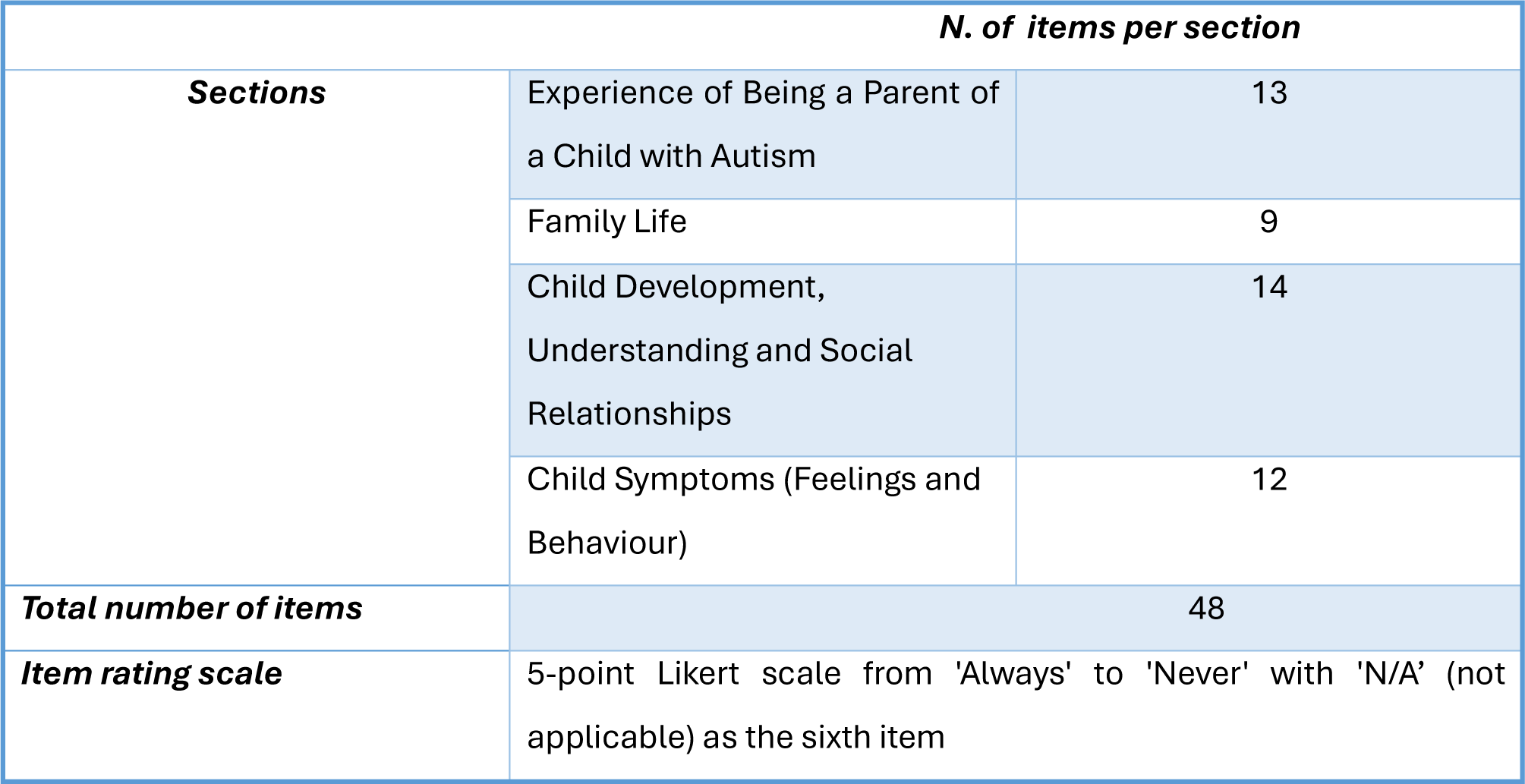
Characteristics of the AFEQ (Leadbitter et al., 2018).

## Methods

Linguistic validation and cross-cultural adaptation to the Italian context were conducted based on Beaton and colleagues’ (2000) guidelines, which provide for the adaptation of individual items, questionnaire instructions and response options. According to Beaton and colleagues (2000), the aim of the cross-cultural adaptation process is to achieve content equivalence between the original material and the translated one; despite what one might intuitively think, the process does not necessarily maintain the psychometric properties of the instrument, such as the validity and reliability at item and/or scale level; therefore, once the translation phase has been completed with the favourable opinion of the committee of experts recruited for the study, it is necessary to carry out further tests on the psychometric properties of the adapted questionnaire.

Following the consensus obtained by the authors of the original instrument and following the recommendations of Polit and colleagues (2007) and Polit and Beck (2006), a second group of selected experts determines the content validity of the adapted instrument through the calculation of the Content Validity Index (CVI), according to the indications for each item (I-CVI) and for the scale as a whole (S-CVI). Figure 1 below describes the steps of the linguistic validation.

**Figure 1.**
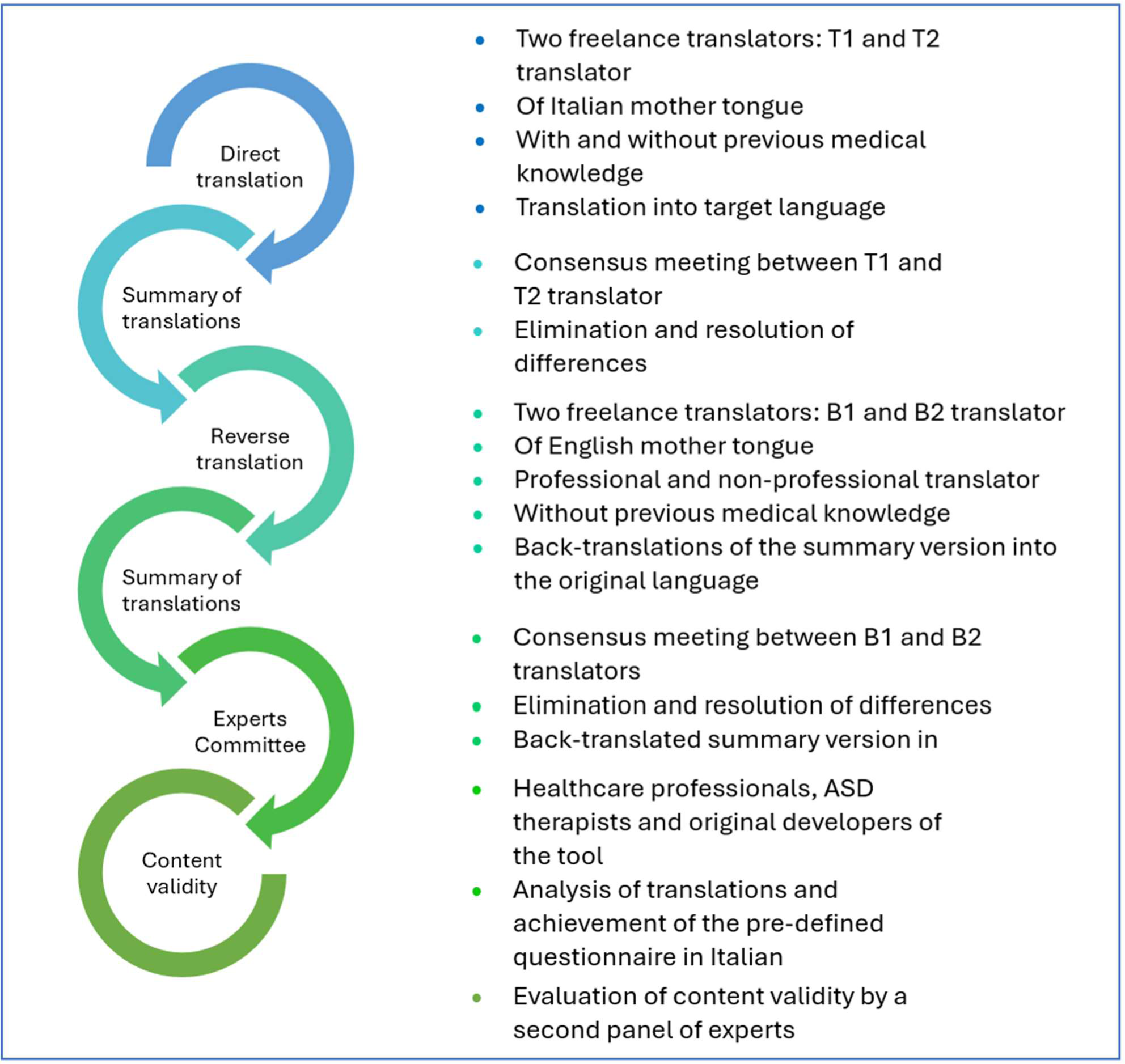
Stages of the linguistic validation and cultural adaptation of the AFEQ to the Italian context.

### Direct, reverse and synthesis translation

The first phase of the research involved the translation of the questionnaire from the original language into Italian, in the period June-July 2023. Two Italian mother-tongue translators were involved, who do not work in this field by profession but are linguistically competent in both English and the target language. The first translator, who has decades of experience in the medical field, both clinical and research, focused on producing equivalence from a thematic point of view, while the second, who has a degree in foreign languages and literature and is not involved in the subject matter (what Beaton and colleagues (2000) call a ‘naive translator’) focused on semantics, ensuring that the translation reflected the language used in everyday life, which is neither technical nor restricted to the healthcare field. The two translators worked independently, producing two separate translations (T1 & T2).

At the same time, the two translators met and produced a synthesis (T-12) of their respective translations by reflecting on and discussing different points of view and arriving at a syntactically and semantically unified version. Subsequently, two native English-speaking translators were recruited for back-translation or reverse translation from Italian into English; the two translators are one a professional working in the tourism sector, the other a professional translator, both with no knowledge of the health-rehabilitation field. Each worked on the T-12 questionnaire which was translated independently and in blind; again, both translators, who had no previous knowledge of the subject and were unfamiliar with the objectives of the study, produced two separate back-translations (B1 and B2).

### Approval by the Committee of Experts

In this phase, the original questionnaire and each translation obtained in the previous phases were made available to an interdisciplinary committee of experts; the latter consisted of Research Fellow Kathy Leadbitter, author of the original questionnaire and professor in the Faculty of Psychology and Mental Health at the University of Manchester, Dr Francesca Bonetti, lecturer in the discipline of Physiotherapy Rehabilitation Science 2 at the University of Rome ‘Tor Vergata’ and researcher in the validation of some scales and questionnaires in the field of physiotherapy, and the four translators (T1,T2,B1,B4) involved in this process. The objective achieved by the committee was to examine and compare the translations, confirming the direct version as the pre-final version in Italian.

### Assessment of Content Validity: The Content Validity Index (CVI)

Although there are different definitions of the construct of content validity in literature, according to Haynes and colleagues (1995) content validity represents "the degree to which the items in an assessment instrument are relevant and representative of the target construct for a particular assessment purpose". Fain (2004) adds to Haynes and colleagues’ (1995) definition that content validity "is determined by a panel of experts carefully evaluating all items and the appropriateness of the instrument for a given population and for measuring a variable or concept". Between September and October 2023, based on a criterion of educational and professional convenience, 7 professionals with expertise in autism collaborated to determine the content validity of the AFEQ in its Italian version.

For each item of the questionnaire, each expert, based on Polit and Beck (2006), assigned a score using a 4-point scale with the following scores:

1. Not relevant
2. Partly relevant
3. Somewhat relevant
4. Very relevant

Following the criteria of item acceptability incorporating the standard error of proportion developed by Lynn (1986) and described by Polit and Beck (2006), in the case of six (or more) experts, the value of the I-CVI for each item was set at greater than or equal to 0.78. For the calculation of the S-CVI, Polit and Beck (2006) describe two variants; for this reason, both are calculated: in the case of more than five experts, whereas for the S-CVI/UA the acceptability cut-off is greater than or equal to 0.80 for the S-CVI/AVE it is 0.90.

Below are the formulas used to calculate both parameters:

- 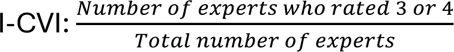
- 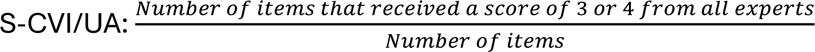
- 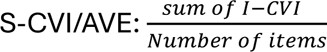

## Results

### Results of direct translation and synthesis

For the direct translation process, two translators (T1 non-health expert, T2 expert), both native speakers of Italian but with sufficient knowledge of English to grasp the different nuances, were involved, who independently produced a translation. Subsequently, the two translations were compared and discrepancies in the translation process smoothed out in a discussion between the translators. The difficulty, but also the crucial aspect, of this translation phase turned out to be being able to find the right compromise between a more technical translation and a more ecological one, comprehensible to the target audience of the questionnaire, i.e. the parents of children with ASD; specifically, among the main items that engaged the discussion between the two translators we mention some examples:

- *Item* 4: “*I am confident that I understand my child’s level of development*” translated by T1 as "I am certain that I understand my child’s level of development" and by T2 as "I am confident that I understand my child’s level of development"; the term "confident" according to the WordReference.com (WordReference English-Italian Dictionary©; 2023) can be translated as "secure, certain, convinced"; although the adjective "confidente" belongs to the Italian language, it is almost exclusively a prerogative of the literary language; with the influence of the English language it is spreading into everyday language, having the meaning of "confident" (Eliseo, 2017). Since the term ‘confidente’ although spreading in recent years may not be easily understood, it was decided to use the translation of T1.
- *Item* 22: “*My child has fussy eating that makes it diflicult to go away for a break*” translated by T1 as ‘My child has such particular tastes in food that it is difficult to go away for a break’ and by T2 as ‘My child has a very particular way of eating that makes it difficult even to take a short break’; in this item, the term that caused the most difficulty in translation is ‘fussy eating’; the latter is translated according to the WordReference Online Dictionary. com (WordReference English-Italian Dictionary©; 2023) with "person with difficult tastes"; children with autism often present eating problems, such as a preference for the same foods or different eating behaviour than their typically developing peers (Diolordi et al, 2014). As food selectivity is a major problem for many children with ASD (Cermark et al., 2010), of the two translations, the one proposed by T1 that is most in line with the need to investigate the presence of this issue in the child was chosen.
- *Item* 24: “*My child can spontaneously begin communication with me”* translated by T1 as "My child spontaneously communicates with me" and by T2 as "My child is able to spontaneously begin to communicate with me"; in this case the translation of T2 was agreed upon as the former would have lost the word "begin", an indication of assertiveness on the part of the child. The same principle was followed for item 25.
- *Item* 30: “*My child has repetitive behaviour and sensory interests that make it diflicult to go on an outing”* translated by T1 as ‘My child has repetitive behaviour and sensory interests that make going out difficult’ and by T2 as ‘My child has repetitive behaviour and sensory interests that make going out difficult’; of the two translations, the second was selected as ‘repetitive behaviour and sensory interests’ is a sentence containing technical terms that are shared in the scientific field and used in clinical practice, including in interviews with parents of children with ASD.

Overall, an effort was made by both translators to make the translated questionnaire as faithful as possible to the original, both in terms of clarity, as the target language (Italian) is more complex and verbose than the source language (English), and bearing in mind the purpose of the questionnaire, namely, to investigate the experience of families with autistic children.

### Results of reverse translation and synthesis

The reverse translations were conducted by two independent, native English-speaking translators (B1, B2), neither of whom had experience or knowledge of either the objectives of the study or the healthcare field, starting with the final T1-2 translation into English, the language of the original questionnaire. The version of B2 was much closer to the original than that of B1 in terms of both lexical and structural choices (the translator in charge is a professional translator and has worked on the translation of questionnaires prior to that of the present study). In general, since the two translations are very similar to each other (e.g. in B1 "I have coping strategies to help my child" and in B2 "I have coping mechanisms to help my child") the B2 translation was chosen as it is almost superimposable on the original except for item number 36 which in the original reads "I have to go with my child to supervise play with other children" while in the B2 version it is "I must go with my child to supervise play with other children"; the two sentences differ only for the verb used, "have to" in the original and "must" in the B2 translation, both used in English to express an obligation/duty (WordReference English- Italian Dictionary©; 2023).

### The Committee of Experts’ Conclusions

The revision process involved individual work, in which each expert individually analysed the material produced in the previous steps. In the case of the present study, no subsequent meeting was necessary, as all members of the expert group agreed on the translated version T1-2 as the only shared tool.

### Content Validation Outcomes

Table 2 below describes the number and main characteristics of the experts who took part in the content validity assessment of the ‘Autistic Family Experience Questionnaire (AFEQ)’.

**Table 2.** Characteristics and size of the expert group.

| EXPERTS | PROFESSIONAL EXPERIENCE | WORK POSITION | PROFESSIONAL FIELD |
| --- | --- | --- | --- |
| 1 | 5 years | Childhood Neuro and Psychomotricity | Assessment and treatment of neurodevelopmental and learning disorders |
| 2 | 10 years | Childhood Neuro and Psychomotricity | Assessment and treatment of neurodevelopmental and learning disorders |
| 3 | 5 years | Childhood Neuro and Psychomotricity | Assessment and treatment of neurodevelopmental and learning disorders |
| 4 | 19 anni | Speech therapist | Assessment and treatment of neurodevelopmental, speech, learning and communication disorders |
| 5 | 15 years | Psicologa dello Sviluppo e dell'Educazione, Terapista della | Psychotherapeutic treatment, assessment and treatment of |
|  |  | Neuro e Psicomotricità dell'età Evolutiva | neurodevelopmental and learning disorders |
| **6** | Over 9 years | Psychologist, Psychotherapist in training - RBT Certified Behavioural Technician | Psychotherapeutic and behavioural treatment |
| **7** | 20 years | Child neuropsychiatrist | Diagnosis and treatment of neurological and psychiatric diseases with onset in the 0-18 age group |

Analysing the I-CVI, all items exceeded the expected cut-off of ≥ 0.78 except item number 14 ‘Family life is a battle’ which obtained an I-CVI value of 0.6. The scores provided by each expert are shown in Table 3.

**Table 3.**
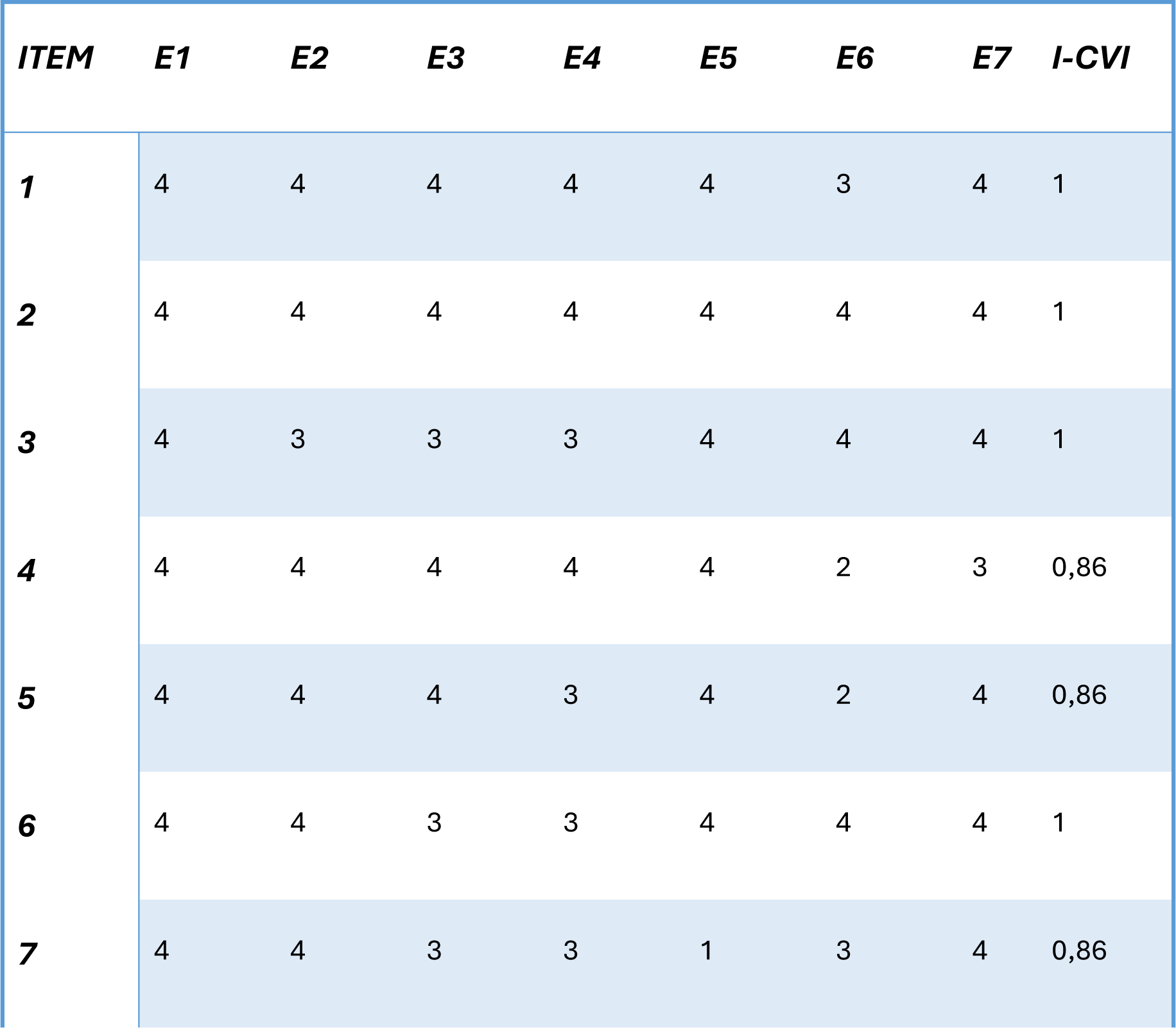
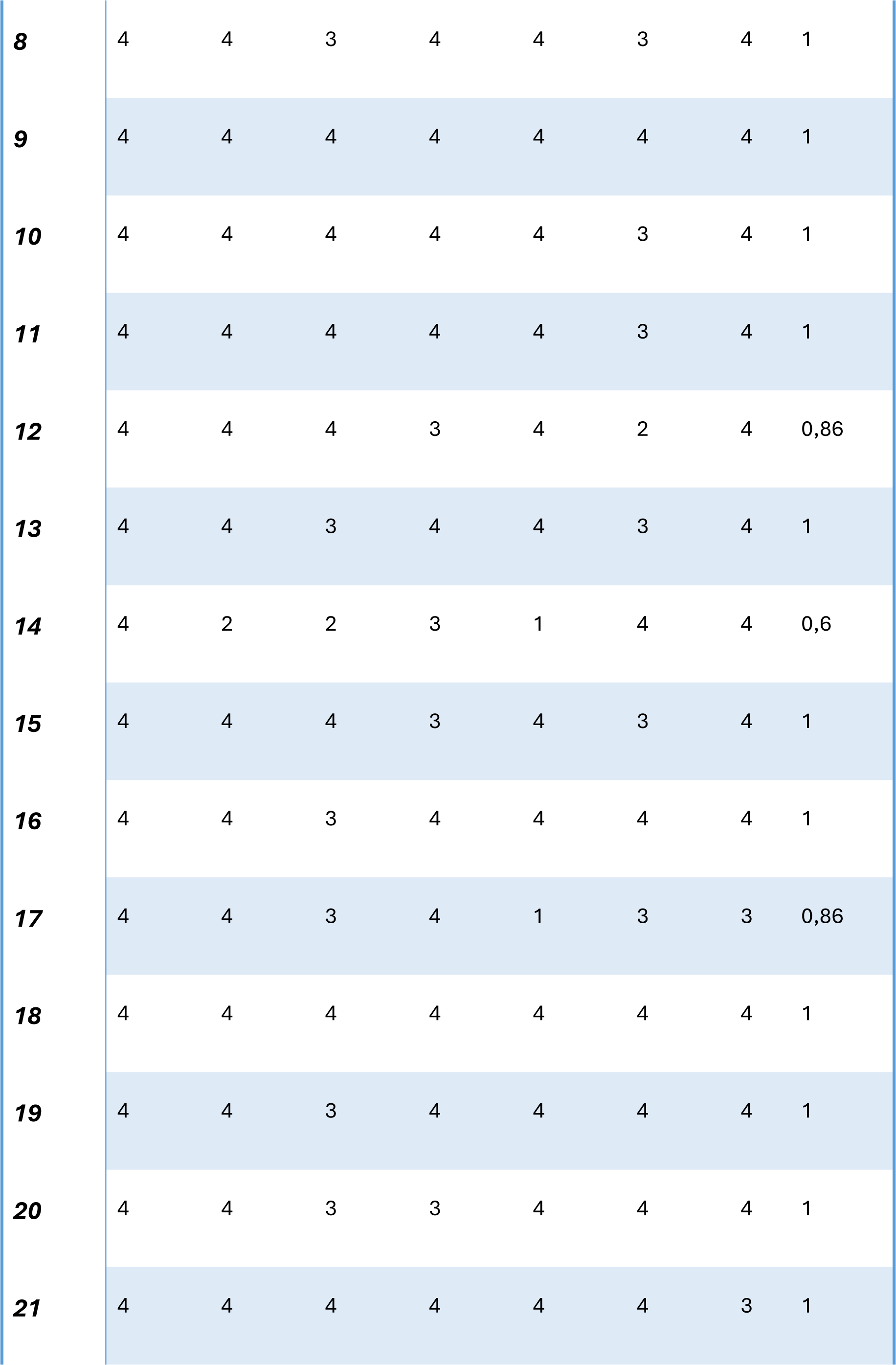
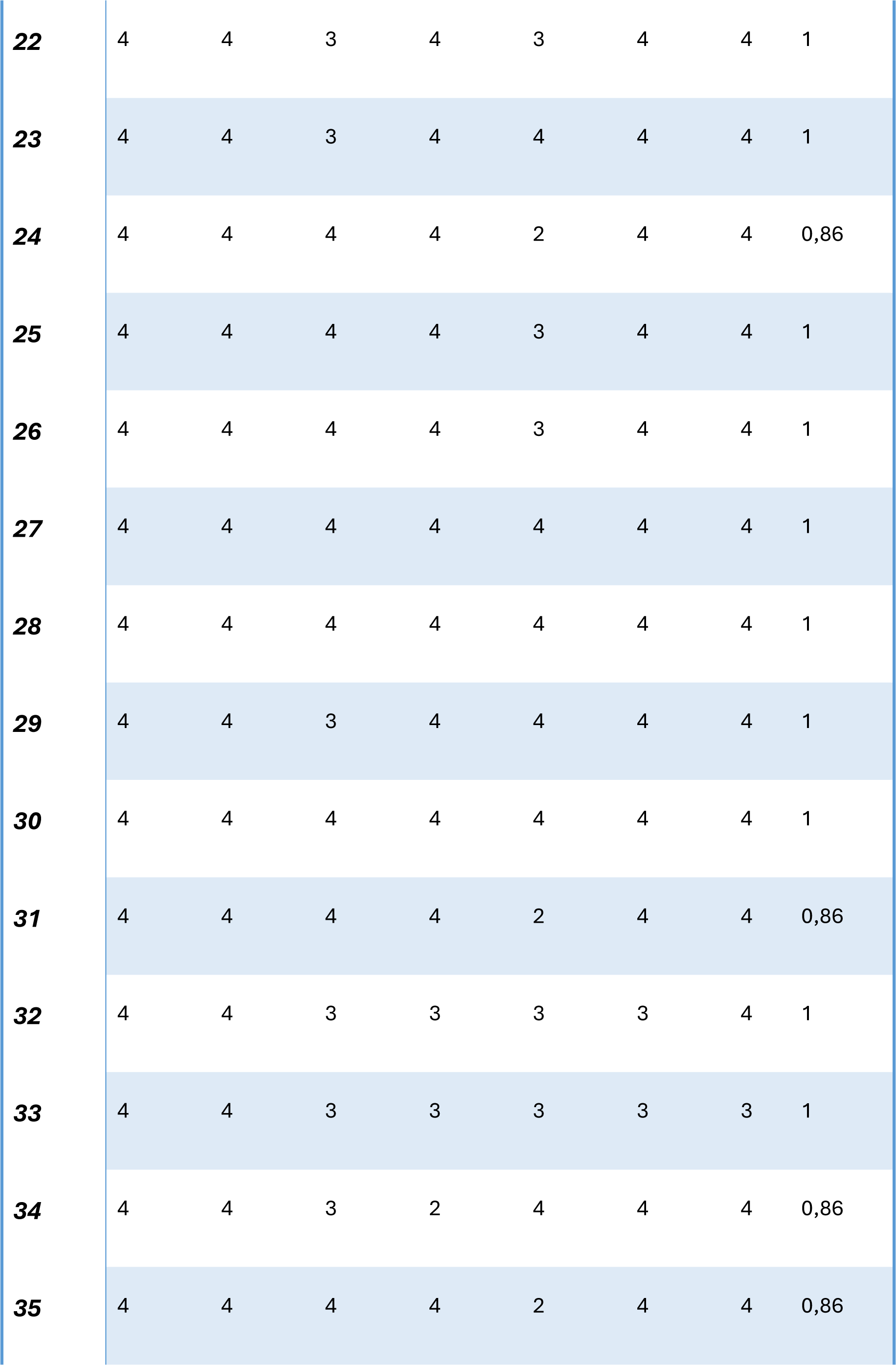
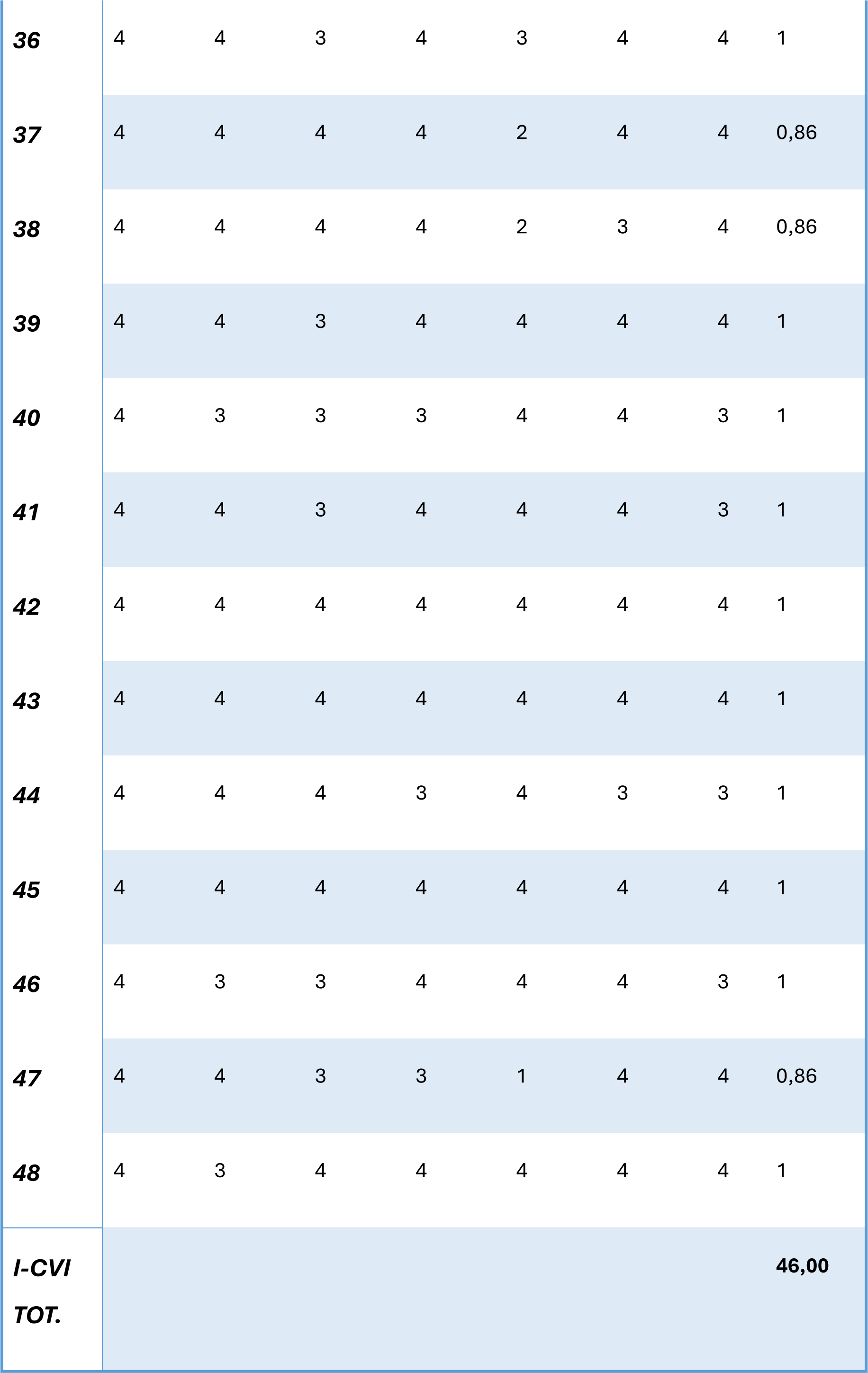
Description of the I-CVI values obtained.

The S-CVI, as shown in Table 4, reached and exceeded the cut-off if obtained through the S-CVI/Ave calculation mode, whereas with the S-CVI/UA the value is 0.73 and is slightly below the cut-off.

**Table 4.**
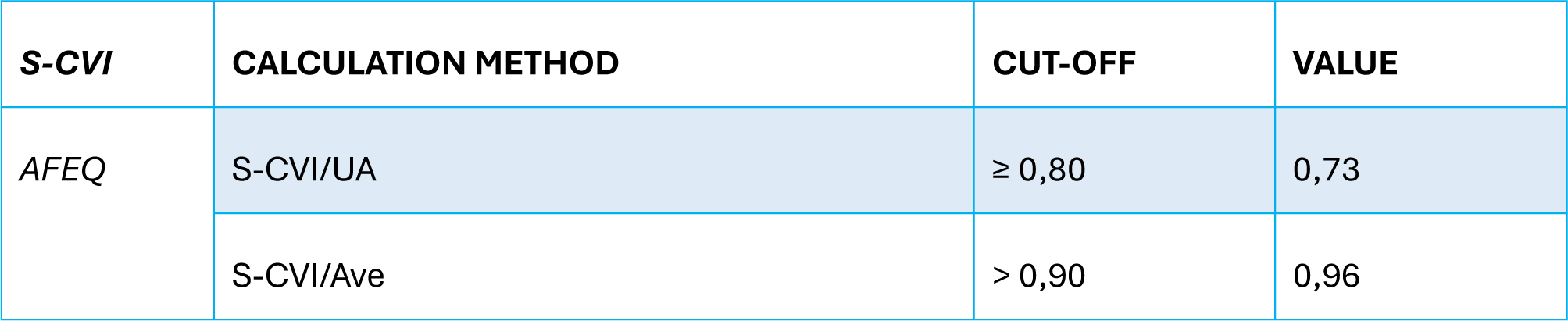
Description of the obtained S-CVI values.

| S-CVI | CALCULATION METHOD | CUT-OFF | VALUE |
| --- | --- | --- | --- |
| AFEQ | S-CVI/UA | ≥ 0,80 | 0,73 |
|  | S-CVI/Ave | > 0,90 | 0,96 |

## Discussion

The aim of the present study is to develop an Italian version of the AFEQ questionnaire, which can respond to the need typical of clinical practice to gather information on the family experience of parents of children with autism of pre-school age in order to capture significant changes subsequent to the rehabilitation treatment of the child and the family unit. The use of the questionnaire could therefore prove to be very useful at the clinical level for guiding diagnostic and rehabilitation choices for the child, taking into consideration the well-being of the parents and the entire family system, an aspect that particularly in the context of interventions aimed at young children is strongly recommended to be evaluated (Hastings et al., 2014; Payakachat et al. 2012; Tint and Weiss, 2016).

The methodology used makes it possible to obtain a version of the tool translated into Italian, which was not yet present in the literature prior to this study. Having rigorously followed what has been indicated by various authors (Beaton et al., 2000; Polit et al., 2007; Polit and Beck, 2006) is a strength of the present study. Involving two translators in the forward translation avoided more bias than would have been achieved by a single translation and enriched the process of producing the T-12 synthesis. The involvement of a figure from outside the medical context on the one hand and a figure familiar with the technical-scientific terminology in the health sector on the other, both with profound knowledge of the English language, made it possible to grasp lexical nuances, choose the most appropriate option for the instrument and at the same time empathise with the point of view of a parent.

The effectiveness of back-translation lies in the involvement of two mother-tongue experts who helped verify the semantic equivalence of the translation (Beaton et al., 2000): the back-translators worked completely independently, producing two separate versions, one of which overlapped with the original except for one item, demonstrating that despite the changes made in the process of adaptation from English to Italian, the meaning of the original text was maintained and thus the authors’ intent. The final version, once analysed, was submitted to a panel of experts for evaluation (Beaton et al., 2000). The panel of experts constitutes one of the greatest strengths of the work: it ensured linguistic validity.

The way it was carried out envisaged a first part of independent revision and a second part of group discussion to allow each expert to reflect on what was requested and then bring his or her own point of view to the common synthesis, resolving any problems encountered. In the case of the present study, a subsequent meeting was not necessary, as all members of the expert group, after reflecting on what was requested, agreed on the T1-2 translated version as the only shared tool, without the need for any improvements. A second group of experts was then recruited, selected so as to include professionals with clinical and rehabilitation experience in the field of Autism Spectrum Disorder and who therefore deal with families with children with ASD every day in their clinical practice; the task of this group was to attribute relevance scores to each item in order to demonstrate the actual content validity of the questionnaire from a qualitative point of view (Polit et al., 2007). The Italian version of the AFEQ shows I-CVI values ranging between 0.83 and 1 except for item number 14 ("Family life is a battle") whose value is 0.6; despite the score being below the cut-off, the members of the panel of experts, after a suitable comparison, decided to keep it as more than half of the experts consider it a relevant or extremely relevant item. The questionnaire also presents a value of S-CVI/UA of 0.73 and of S-CVI-Ave of 0.96 showing in general a very positive judgement by the experts on the items of the scale in terms of relevance with respect to the constructs investigated.

### Limits of the study and prospects

Despite the promising results described above, the study is not without its limitations. The main limitation of the work presented is methodological: as the study is still ongoing, it has not yet been possible to carry out a test of the adapted version that coincides with the last phase (the pretest phase) of the adaptation process described by Beaton and colleagues (2000). The pretest consists of administering the obtained version in the target language to a group of subjects or patients (ideally between 30 and 40 people) specific to the reference context. Once each subject has completed the questionnaire, they are interviewed to verify the possible presence of a high percentage of missing items or single answers, a symptom of a reduced understanding of the questions and therefore of a poor equivalence between the original and the adapted version. Despite this, the main objective of this phase of the adaptation process is to provide a certain measure of the content validity of the instrument (Beaton et al., 2000) which, although not on the parents to whom the questionnaire is addressed (External or façade), was evaluated by a selected group of experts through the calculation of the I-CVI and S-CVI indices. The work described in the previous paragraphs therefore represents a first stage in the Italian validation process of the instrument proposed by Leadbitter and colleagues (2018), which will need to be continued with validation on many subjects. The present study intends to present itself as a methodologically robust basis from which to build a path that, through further studies, will lead to making the AFEQ questionnaire available in the Italian context. Since Europe is a multilingual continent, translations play a fundamental role in comparing the results of different studies and establishing their replicability in other countries. The study therefore aims to provide researchers and clinicians with a new tool to assess the rehabilitation outcomes expected and perceived as a priority by parents of pre-school children with ASD, as well as providing information on the quality of life and well-being of the entire family unit. Being a questionnaire aimed at parents, the study offers the benefits described above at zero risk. The adaptation of a questionnaire intended for a new linguistic context is undoubtedly a time-consuming and costly process, however, to date it is considered the best method to maintain the psychometric properties of the instrument considered (Beaton et al., 2000). Through the process of translation and adaptation, it is possible to compare data collected in studies involving different countries or to avoid selection biases; the latter are defined by Tripepi and colleagues (2010) as "errors in the selection of study participants" such as, for example, the need to exclude some subjects not because they did not possess the inclusion criteria to participate in the study, but because they were unable to fill out a form in English, given the lack of translated versions of the questionnaire (Beaton et al., 2000). The validated Italian version of the questionnaire by Leadbitter and colleagues (2018) can therefore be used by all those who deal with Autism Spectrum Disorder, from child neuropsychiatrists to therapists for monitoring the outcomes of a rehabilitation intervention, in order to obtain useful information for designing interventions directed at children with ASD and their families.

## Data Availability

All data produced in the present study are available upon reasonable request to the authors.

## Conflicts of interest

The authors declare no conflicts of interest.

## Notes

### Competing Interest Statement

The authors have declared no competing interest.

### Funding Statement

This study did not receive any funding

### Author Declarations

Istituto Leonarda Vaccari' Ethics committee gave ethical approval for this work

